# Opportunities for cost reduction of current first-line WHO-recommended oral antiretroviral therapy: replacing tenofovir disoproxil fumarate with tenofovir alafenamide

**DOI:** 10.64898/2026.03.12.26348214

**Authors:** Lise Jamieson, Willem Daniel Francois Venter, Gesine Meyer-Rath

**Author notes:** **Corresponding author:** Lise Jamieson.

## Abstract

**Introduction:** Dolutegravir-based first-line antiretroviral therapy (tenofovir disoproxil fumarate, lamivudine, and dolutegravir; TLD) has delivered substantial clinical and public health benefits. However, sharply decreasing funding for HIV programmes necessitates cost reduction within current treatment guidelines. We evaluated whether replacing tenofovir disoproxil fumarate with tenofovir alafenamide (TAFLD), a drug with equivalent effectiveness and side effect profile, could reduce HIV treatment costs in South Africa.

**Methods:** We conducted a budget-impact analysis over 2026-2030 from the provider-perspective. The cost of antiretroviral treatment (ART) provision with either TLD or TAFLD was estimated using ingredients-based costing, including the cost of drugs, laboratory monitoring, staff, consumables, equipment and overheads. Costs are reported in 2025 USD, are undiscounted and not inflated. Population estimates for adults on first-line therapy were derived from Thembisa 4.8. We modelled a phased transition from TLD to TAFLD over two years, and explored sensitivity to TAFLD price variation (±15%) and inclusion of creatinine monitoring.

**Results:** TAFLD reduced per-patient annual costs by 4-5% compared with TLD (from US$178 to US$169, and US$287 to US$277, for first and follow-up years, respectively). At full replacement, total programme savings were approximately US$54 million per year (-5%). Even with continued creatinine monitoring, TAFLD remained cost-saving, reducing annual costs by around 4%. Savings increased to 8% if TAFLD prices were 15% lower than base-case assumptions.

**Conclusions:** Replacing TDF with TAF in first-line antiretroviral therapy could generate meaningful cost savings for South Africa with minimal programme disruption. While long-term metabolic effects require consideration, TAFLD represents a feasible interim cost-reduction strategy while awaiting next-generation HIV therapies.

## Introduction

Current World Health Organization (WHO) guidelines have recommended a combination of dolutegravir and tenofovir disoproxil fumarate (TDF) plus either lamivudine or emtricitabine for all people living with HIV since 2019, for reasons of improved persistence, safety and resistance over previous regimens [1,2]. The introduction of this regimen has been a public health success, with an estimated 29 million people, or 91% of the global total of those living with HIV, on a combination of TDF, lamivudine and dolutegravir (TLD) by the end of 2022, at an average cost of US$34 per patient course per year in low- and middle income countries (LMIC) [3]. The cost of efavirenz-based annual therapy, now replaced by TLD in a country like South Africa with over 6 million people on therapy in 2025, in 2017 was US$110/year, although other low-and middle-income countries paid less due to centralized procurement agreements. Our previous modelling work has demonstrated substantial benefits in terms of life-years saved and HIV transmission averted by the transition to TLD [4].

However, with health budgets under severe strain, further exacerbated after recent funding cuts from the United States Government (USG), as well as the continued evolution of antiretroviral therapy, efforts to interrogate opportunities for savings are pivotal. Alternatives exist to the individual components of TLD, and several long-acting agents that might replace the regimen altogether are under investigation, but are unlikely to be a substitute in the near future for TLD for cost, resistance, patent, and other reasons [5,6].

But can individual drugs within TLD be substituted or even removed? The immediate analogue substitute for dolutegravir, bictegravir, brings no added benefit, has less evidence in pregnancy and tuberculosis cases, less experience within large-scale programmes, and is not available in generic form. Lamivudine and emtricitabine are interchangeable, with little price difference, and there is no evidence that they can be removed from the regimen.

Removing TDF altogether from TLD, while offering viral suppression in both clinical trials and real-world high-income country experience, means losing highly effective concomitant hepatitis B treatment, which is common in many places where HIV is prevalent, and where hepatitis B vaccination and screening programmes are often absent. With WHO’s renewed attention to hepatitis B, as well as concerns that dual therapies may perform worse in less well-monitored environments, removing TDF appears to be unlikely to become a recommendation.

Finally, an option would be to replace TDF with tenofovir alafenamide (TAF), another tenofovir prodrug, also available as a generic, although not as widely as TLD. TAF is associated with more weight gain and a less favourable lipid profile when compared with TDF [7]; however, TAF also has a more favourable renal and bone profile, which has led to it being recommended over TDF in most high-income countries [8,9]. Which tenofovir prodrug has a better safety profile is the subject of debate, but both have equivalent virological suppression and hepatitis B coverage, and both medications are very well tolerated [10,11].

TAF also represents several opportunities to lower antiretroviral costs, if replacing TDF. TAF is available at a lower dose (25mg vs TDF 300mg), resulting in possible lower manufacturing costs (see communication with generic manufacturers below) and a smaller pill size, hence reducing packaging requirements. In addition, TALFD requires less active pharmaceutical ingredient (API) due to the lower dose, which could reduce the environmental impact associated with production, distribution, and drug excretion, leading to a lower carbon footprint and cumulative energy demand per patient year. There is speculation that due to the better renal profile [11], creatinine monitoring, currently recommended for TDF, could be removed if TAF was introduced into programmes. Finally, switching clients from TDF to TAF would be swift, as no programme preparation would be required, unlike the lengthy transition from stavudine to tenofovir, or from dolutegravir to efavirenz, which required laboratory assessments, as well as patient and health worker preparation.

We estimated what the South African programme would save annually if TLD were to be replaced by its equivalent of TAF, lamivudine and dolutegravir (TAFLD), with and without creatinine monitoring.

## Methods

### Cost estimates

Cost inputs were sourced in 2025, based on prevailing prices at that time, and the subsequent modelling and analysis were completed in 2026. A budget impact analysis was conducted from the perspective of the provider, the South African government, over a 5-year period from 2026 to 2030. In South Africa, a 5-year timeframe is generally preferred to assess mid-term fiscal implications.

We used ingredients-based costing to estimate the cost of antiretroviral (ARV) treatment provision per patient-year at a general primary health clinic (PHC) for both TLD and TAFLD arms. Resource use in terms of number of clinic visits, staff time and cadre required, type and quantity of non-ARV drugs, equipment and other overheads, including space and utilities, were informed by an economic evaluation of such a clinic by Long et al [12]. Laboratory testing (viral load, CD4 count and creatinine) and ARV drug quantities were assumed as per South African treatment guidelines [13]. Unit costs were sourced from government sources: staff salaries were based on the Department of Public Service and Administration [14], TLD and other non-ARV drug prices on the Master Health Product List [15], laboratory costs on the National Health Laboratory Service (personal communication with NHLS). Clinic-level overhead costs sourced from Long et al were inflated to 2025 prices using the Consumer Price Index estimates [16]. The price of TAFLD was informed by discussions with two generic manufacturers, both of which confirmed that the price of TAFLD could be set at US$3.64 per 28 tablets. In contrast, TLD is currently bought from various suppliers, by the South African government, for a weighted average price of US$4.17 for 28 tablets (weighted by quantity awarded across suppliers) [15]. In sensitivity analysis, we included the costs of creatine separately in the event that this does get included in initial TAFLD guidelines, and additionally ranged the unit cost of TAFLD by ±15%.

All costs are presented in 2025 US$, undiscounted, and converted using the average exchange rate for January-May 2025 of ZAR 18.54 per US$1 [17].

### Population estimates

We used Thembisa 4.8 to estimate the number of men and women aged 15+ years on first-line HIV treatment between 2026 and 2030 [18], assuming the current trajectory of treatment coverage. Thembisa reports the number of PLHIV aged over 15 years starting ART mid-year points, currently on ART, and on second line ART. Laboratory monitoring and number of clinic visits differ between the first year of ART and follow-up years thereafter, resulting in different costs between these periods. Patients on second line ART were not considered for TAFLD.

We assumed that all PLHIV currently on TLD would switch over to TAFLD over the course of 2 years, with 50% having moved over by the end of 2026 and 100% by end-2027. The pace of transition for TLD to TAFLD was modelled on transition from TEE to TLD between 2020-2022. That transition happened slower than anticipated, for complex reasons ranging from concerns around perceived side effects to residual unused stock, and while a transition from TLD to TAFLD should be simpler, we have assumed that similar unanticipated system complexities may slow the changeover.

### Ethics and consent

This study was based on mathematical modelling using publicly available, aggregate data and did not involve human participants. Therefore, ethics approval was not required, and informed consent was not applicable

## Results

The full-service cost of TLD per patient-year is estimated at $287 (first year of ART) and $178 (follow-up years). In comparison, TAFLD in the first year of ART is estimated to cost $277 per patient year, while follow-up years are estimated to cost $169 per patient year, a 4-5% reduction in costs. While total cost of TLD provision is set to range between US$1,063 and US$1,163 million over the next 5 years, if HIV treatment coverage remains at the current trajectory, TAFLD implementation can reduce the cost of first-line treatment to between US$1,031 and US$1,080 million by 5% at full replacement, a saving of approximately US$54 million annually, or 5%. If the price of TAFLD were reduced by 15%, full replacement with TAFLD could lower annual first-line treatment costs by 8%. Conversely, a 15% increase of the price of TAFLD would make it slightly more expensive (+0.1%) than the current TLD regimen.

If creatinine testing were to remain as part of monitoring as it is with TLD, it would cost of US$12 million per year for current first-line patients; however, this cost is minimal, compared to the overall cost of treatment, and even with it added, TAFLD would still reduce the cost of first-line treatment by 4% annually (total costs between US$1,043 and US$1,093 million) at full replacement.

**Table 1.** Cost of ART provision for TLD vs TAFLD over 2026-2030.

|  | 2026 | 2027 | 2028 | 2029 | 2030 |
| --- | --- | --- | --- | --- | --- |
| **Number of adults on first-line treatment (millions)** |  |  |  |  |  |
| Number on TLD | 2.92 | 0.00 | 0.00 | 0.00 | 0.00 |
| Number on TAFLD | 2.92 | 5.97 | 6.09 | 6.19 | 6.28 |
| **Total** | **5.85** | **5.97** | **6.09** | **6.19** | **6.28** |
| **Total cost of TLD provision (USD, millions)** |  |  |  |  |  |
| ARV costs | 314 | 320 | 327 | 332 | 337 |
| Cost of provision* | 749 | 763 | 777 | 789 | 799 |
| **Total** | **1,063** | **1,084** | **1,104** | **1,121** | **1,136** |
| **Total cost of TAFLD provision – without additional creatinine testing (USD, millions)** |  |  |  |  |  |
| ARV costs | 294 | 280 | 285 | 290 | 294 |
| Cost of provision* | 743 | 752 | 765 | 777 | 786 |
| **Total (% change over TLD)** | **1,037 (-2%)** | **1,031 (-5%)** | **1,050 (-5%)** | **1,067 (-5%)** | **1,080 (-5%)** |
| **Total cost of TAFLD provision – with creatinine testing (USD, millions)** |  |  |  |  |  |
| ARV costs | 294 | 280 | 285 | 290 | 294 |
| Cost of provision* | 749 | 763 | 777 | 789 | 799 |
| **Total (% change over TLD)** | **1,043 (-2%)** | **1,043 (-4%)** | **1,062 (-4%)** | **1,079 (-4%)** | **1,093 (-4%)** |

\*Provision costs include staff, laboratory testing, non-ARV drugs, equipment and overheads

## Discussion

Our modeling suggests that under a full-scale roll-out of TAFLD at its current estimated generic price, the South African programme could save about US$54 million per year at full replacement, or $9-11 per patient-year, with a move to TAF from TDF. In addition, there could be a slight simplification to the programme by removing creatinine monitoring, given the more favourable renal profile that TAF provides over TDF, saving an additional US$12 million annually. While these are small proportional savings, at 5% of the total cost of first-line ART, they are relevant especially in the current climate of severe reductions in programme support by the US government.

The transition in this case is simpler than previous regimen changes. In contrast to the move from stavudine to tenofovir, this transition does not require any additional laboratory monitoring. Further, in contrast to the move from efavirenz to dolutegravir, the move will not require additional patient counselling, as the side effect profile is likely similar. In both prior examples, substantial health worker training was required, and little additional training would be required for this change.

Our budget impact analysis does not factor in the costs associated with potential unknown sequelae such as weight gain and lipid changes. However, and similarly, it does not factor in the bone and renal changes associated with staying with TDF on these regimens. Balancing the complex and myriad obesity-linked health consequences against concerns about rising renal disease and loss of bone density, in a situation where African data on these long-term consequences is near-absent, is an impossible task. Weight gain in people with HIV has emerged as a major concern and is associated with substantial costs to the health sector [19]. However, a more comprehensive long-term approach to weight gain for people living with HIV, and, indeed, the country, is required [20]. Moreover, it is likely that TLD, TAFLD, and current oral first-line therapies, judging by the speed of prior antiretroviral evolution, will be replaced by next generations of long-acting agents, although this appears unlikely for the next decade [5,6].

Beyond this, limitations to our analysis include the following: Our model presupposes the entry of several large generic competitive manufacturers, with adequate notice to generate the necessary manufacturing capacity. At the last entry of TLD, the South African government insisted on six major manufacturers being part of the tender, as a means of de-risking supply. Our model also assumes our information regarding prices from the generic companies we canvassed is correct. We assume that TAF API supply will be as plentiful as for TDF. We assumed that guideline groups and National Essential Medicines List Committee (NEMLC), the ministerial advisory body responsible for developing and reviewing the country’s Essential Medicines List (EML) for the public health sector, might prioritize cost considerations even in the context of potential concerns over weight gain and lipid changes. Finally, we do not have robust data on the number of people accessing or taking up ART currently, due to the loss of PEPFAR-supported data-systems, impacting on the accuracy of projected savings.

## Conclusions

Cost reduction options on current first-line antiretroviral recommendations are limited, and a transition from TDF to TAF while awaiting the next generation of long-acting agents may yield savings. However, careful consideration must be given to currently unknown long-term costs resulting from a transition to TAF, such as weight gain.

## Authors’ contributions

LJ and WDFV conceptualized the work, and drafted the manuscript. LJ and GMR conceptualised the analytical framework and developed the unit costs. LJ completed the economic analysis. All authors critically reviewed and approved the final manuscript, and share final responsibility for the decision to submit for publication. All authors had access to all data in the study.

## Data Availability Statement

The modelling outputs used are publicly available at https://thembisa.org/downloads

